# Forecasting COVID-19 New Cases Using Transformer Deep Learning Model

**DOI:** 10.1101/2023.11.02.23297976

**Authors:** Saurabh Patil, Parisa Mollaei, Amir Barati Farimani

## Abstract

Making accurate forecasting of COVID-19 cases is essential for healthcare systems, with more than 650 million cases as of 4 January,^1^ making it one of the worst in history. The goal of this research is to improve the precision of COVID-19 case predictions in Russia, India, and Brazil, a transformer-based model was developed. Several researchers have implemented a combination of CNNs and LSTMs, Long Short-Term Memory (LSTMs), and Convolutional Neural Networks (CNNs) to calculate the total number of COVID-19 cases. In this study, an effort was made to improve the correctness of the models by incorporating recent advancements in attention-based models for time-series forecasting. The resulting model was found to perform better than other existing models and showed improved accuracy in forecasting. Using the data from different countries and adapting it to the model will enhance its ability to support the worldwide effort to combat the pandemic by giving more precise projections of cases.

## Introduction

Since late December 2019, there has been a negative influence on social norms and values, The COVID-19 pandemic has had a profound and far-reaching impact on almost every aspect of human life. Since late December 2019, there has been a negative influence on social norms and values, lifestyles, the wealth of countries, and the psychological behavior of people all over the world due to the widespread outbreak of COVID-19.^2^ COVID-19 has caused a global health crisis with millions of people infected and hundreds of thousands of deaths worldwide. The disease has overwhelmed healthcare systems, and many people have had difficulty accessing medical care due to hospital overcrowding and a shortage of personal protective equipment (PPE). As of January 4, 2022, over 650 million individuals globally have experienced the repercussion of the SARS-CoV-2 virus. and there have been 6.5 million deaths related to the virus. ^1^ Recent research has shown that the virus can be spread through droplets expelled during breathing and through contaminated surfaces.^3^ These droplets can be inhaled by people who are nearby or can land on surfaces and objects, where the virus can survive for several hours to several days. As a result, large gatherings are recognized as a means of spreading COVID-19, indicating a strong relationship between human movement and the contagiousness of the virus.^4^ Such studies emphasize the significant role of some strategies such as isolation, social distancing, face covering, and closing public places (schools, restaurants, etc.) in diminishing COVID-19 mortality cases.^3^ Even though implementing these policies could slow down the spread of the virus, it remains highly contagious and can be transmitted through various means. In addition, the evolution of dangerous mutations can potentially accelerate the spread of the virus since the new mutations can escape the immune system.^1-3^ This raises demands for additional healthcare resources, hospital facilities, and supplies all over the world.^5,6^ Many researchers have made tremendous efforts to efficiently prevent the spread of the virus,^7,8^ finding effective treatments for COVID-19,^9-12^ vaccines,^13^ antibody designs,^14^ and understanding mutations and modeling.^15,16^ However, all such approaches not only require significant investments but also are long-term solutions suggesting that they may fail to compete with the speed of the spread of the virus in regard to controlling the pandemic. Hence, reliable forecasting of the trend is essentially required for several reasons. Accurate forecasts are necessary for government and public health organizations to plan and allocate resources effectively. This includes everything from the distribution of personal protective equipment to the allocation of hospital beds and other medical resources. Moreover, accurate forecasting allows government and public health organizations to plan their response to the pandemic, including measures such as lockdowns, quarantines, and vaccination campaigns. Reliable forecasts can also help individuals and organizations manage their own risk by allowing them to make informed decisions about their activities and to plan accordingly. Accurate forecasts are necessary for businesses, financial institutions, and governments to plan their economic response to the pandemic. This includes everything from the allocation of stimulus funding to the planning of economic recovery strategies. Furthermore, accurate forecasts are necessary for the research and development of treatments, vaccines, and other medical interventions. This is because the pace of research and development is often driven by the expected course of the pandemic. Finally, reliable forecasts are critical for maintaining public trust in government and public health organizations. If forecasts are consistently incorrect, it can undermine public confidence in these organizations and their ability to respond effectively to the pandemic. In conclusion, reliable forecasting of the COVID-19 trend is essential for effectively managing the pandemic and its consequences, both in the short term and the long term. It is a crucial tool for planning and decision-making at all levels of society. Predicting the direction of the virus will enable us to tailor and implement appropriate public COVID-19 measures in different locations based on the relevant predictions. However, forecasting COVID-19 future cases is very complex and requires accurate modeling. SIR (Susceptible (S), Infected (I), and Recovered (R)) a mathematical model effectively forecasted early COVID-19 case numbers in Indonesia, highlighting the importance of reliable forecasting tools.^22^ Recently, effective utilization of modern deep learning models has been observed in both diagnosis and predictions (using LSTM, CNN, MAE (Modified auto-encoders), etc).^18-22^ For example, Karaçuha et al.^23^ modeled the COVID-19 data and forecasted case numbers for the next 30 days using a practical method based on Fractional Calculus called Deep Assessment Methodology (DAM). In another study, Koç et al.^24^ provided a deep LSTM network containing multiple layers of LSTM network to forecast the expected infected cases. Li et al.^25^ forecasted disseminations of the COVID-19 virus in various nations utilizing a recurrent neural network (RNN) model with attention named ALeRT-COVID. Kalaga et al.^26^ used DL-RNN models to predict reported, recovered, and fatalities cases in different countries. Their model is built on LSTM cells, RNN, and Gated Recurrent Units (GRUs) to forecast COVID-19 trends. Atlam et al.^27^ used the Deep-Cox-COVID-19 model to forecast significant symptoms influencing COVID-19 treatments which combines Cox regression with autoencoder and forecasts the expectation of living by separating out the most severe cases based on the number of fatalities. These approaches provide useful information for pandemic preparedness plans and the management of health services for patients. These models also facilitate making better decisions about lockdown policies. Although all these models are good at forecasting, the accuracy of these models is still a challenge. The emergence of Attention models in time-series prediction^28^ provides the opportunity for developing more accurate forecasting models. The study’s goal was to develop a Transformer model that could anticipate COVID-19 transmission in India, Brazil, and Russia, which are leading nations with an excessive number of cases. We utilized recent advancements in attention models to enhance the precision of our predictions.^29^ To demonstrate the proficiency of our model, we evaluated its precision against CNN-LSTMs, CNN, and LSTM.

## Methods

### Data preprocessing

Data analyzed in research was from January 1, 2021, to January 1, 2022, gathered from [30], it contains information from 210 nations globally starting from January 22, 2020, till the present.^35^ This study explored data specifically from three countries - Brazil, India, and Russia, which have been significantly impacted by the pandemic. To make the data more consistent, a 7-day average of cases was considered, considering the delay in COVID testing, reporting, and updating the data on the portal. As a result, any days with no reported cases were removed from the data. The study used three factors (stringency index, face covering, and stay-at-home) as predictors in its forecasting model. We standardize the data using StandardScaler from the sklearn^36^ module to provide stability in numerical prediction. The features used to allow better understanding of the trends for the DL models is displayed in Table 1.

**Table 1:**
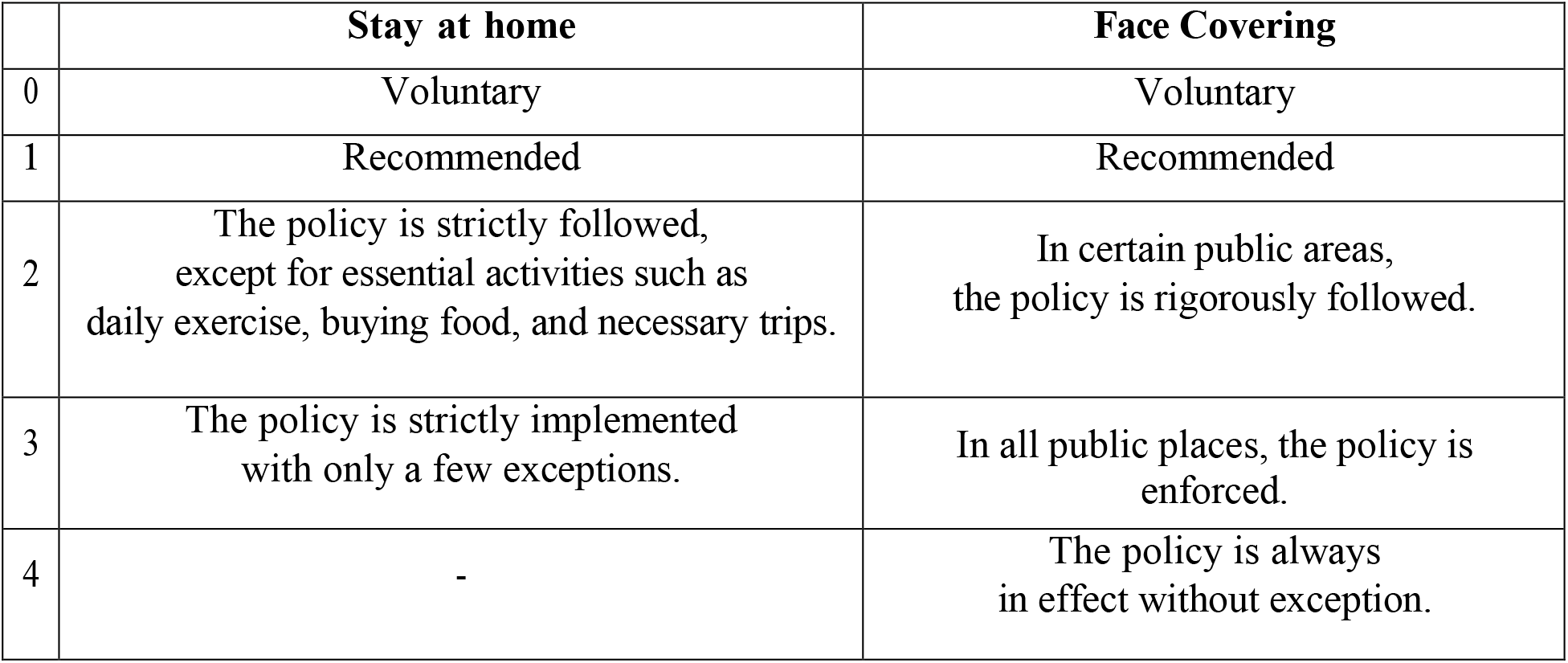
Features used in training data.

|  | **Stay at home** | **Face Covering** |
| --- | --- | --- |
| 0 | Voluntary | Voluntary |
| 1 | Recommended | Recommended |
| 2 | The policy is strictly followed, except for essential activities such as daily exercise, buying food, and necessary trips. | In certain public areas, the policy is rigorously followed. |
| 3 | The policy is strictly implemented with only a few exceptions. | In all public places, the policy is enforced. |
| 4 | - | The policy is always in effect without exception. |

### Models

Following the preprocessing of the data, Transformer^29^ was implemented. The performance of the Transformer was benchmarked against CNN-LSTM, LSTM, and CNN^32^ in order to determine its effectiveness in forecasting COVID-19 cases over the next 4 days. We follow the same parameters for the models that are being benchmarked against as mentioned in [32] and train the models for 300 epochs. Figure 1 describes the transformer architecture and methodology.

**Figure 1:**
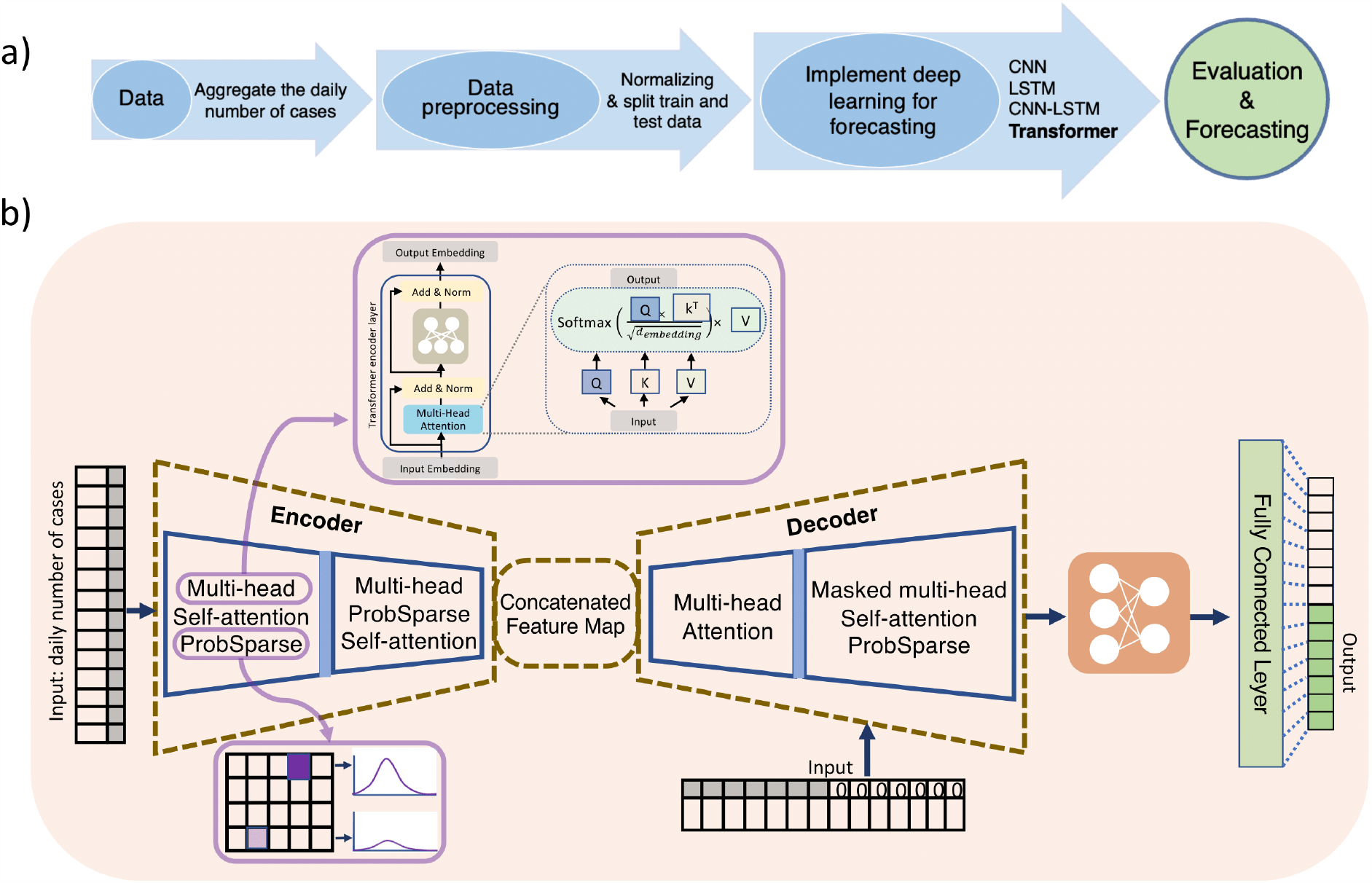
(a) A flowchart was created in this study to represent the methodology used. (b) The Informer architecture is also outlined, with a focus on the use of multi-head prob sparse attention in the encoder to significantly reduce network size and improve efficiency in the decoder by masking target elements and utilizing the feature map for quick output generation.

#### Transformers

A neural network that specializes in processing sequential data, including data that follows a time sequence is a Transformer. Recently, Transformers have been also used in biology for prediction of sequences^37, 38^. It utilizes attention mechanisms to track relationships within the data and to determine the context, allowing it to learn and understand the meaning of the data. The vanilla transformer^28^ is made up of three key elements: the Encoder, the Decoder, and the Attention mechanism. The Encoder serves the purpose of transforming an input data into a compact and continuous vector. The encoded input series includes the number of cases, stringency index, stay-at-home, and face covering, all of which are standardized for numerical stability. The decoder is used to decode the representation vector to generate the target sequence. The decoder receives two inputs: the feature map from the encoder and the lengthy input sequence, with the target elements, masked to zero. The encoder’s feature map is utilized to forecast the target elements. The attention mechanism focuses on the crucial aspect of the input data by learning the connections between components in a sequence automatically. However, using a basic Transformer model for timeseries forecasting had three limitations: 1) the self-attention calculation is too complex, 2) there are memory limitations when stacking layers for long inputs, and 3) there is a decrease in speed when predicting long inputs. To tackle these challenges, the Informer^29^ model was proposed with a novel generative style decoder, Probsparse attention, and a Self-attention distillation operation. The inputs to the encoder are first passed through the convolutional layers for embedding the data into dimension 512, the embedded inputs are passed to the attention block which implements the probsparse attention. The computational requirements for each layer in terms of time and memory utilization is O (L^2^) due to the use of canonical scalar-product in the self-attention mechanism. Alternatively, the probsparse attention only enables each key to focus on the most important queries rather than all of them. This allows the model to compute time-consuming operations for a fraction of queries and reduces the complexity to O (L log L). The probsparse attention uses the below equation, the sparse matrix, referred to as 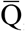, contains the most significant queries.:

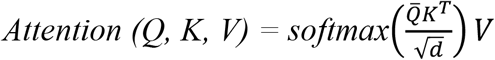

Next, the encoder implements a self-attention distillation where the outputs from the probsparse attention block are passed to the distillation block which consists of a convolutional layer followed by maxpool, where the redundant combinations of value V are removed and only the top features are selected as input to the next attention block. The encoder in Fig 2 is comprised of attention blocks that are divided into halves, reducing in number as the number of layers increases. After the final layers, the feature map is used as the output of the encoder. The decoder structure used in Informer^29^ is the same used as in the Vanilla transformer and it contains multi-head attention and a masked multi-head probsparse attention. The decoder receives the long input sequences, and the target elements are masked with zero, uses the feature map from the encoder, and outputs the target elements in a single forward operation rather than in multiple steps, enhancing the speed of predictions for long sequences.

**Figure 2:**
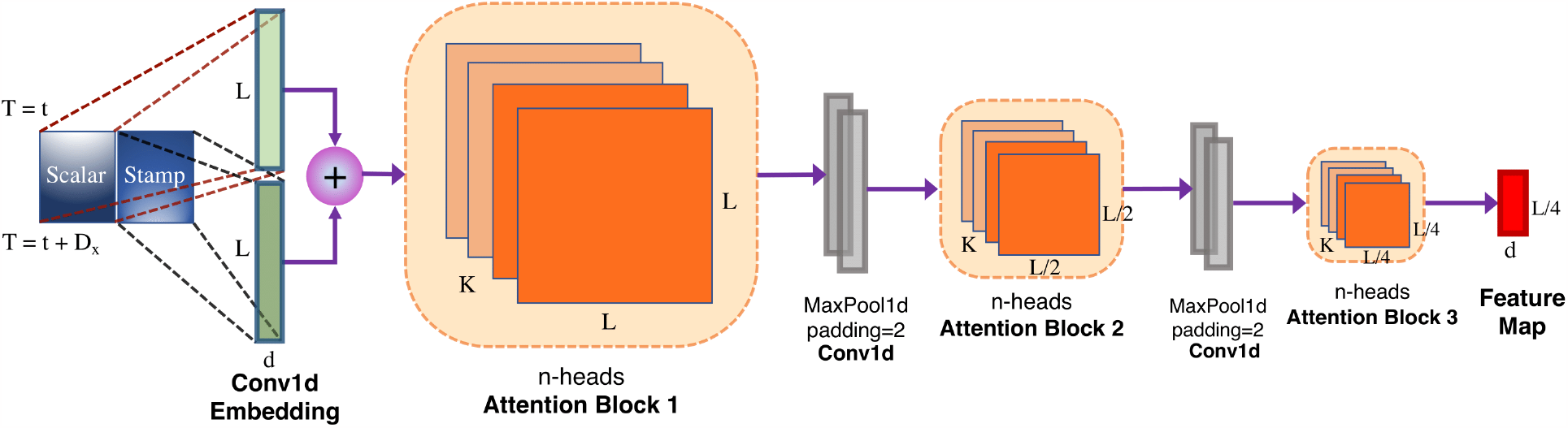
The Encoder architecture used in Informer. The encoder consists of embedding the input data and passing it through the attention block and convolutional layer which distills the redundant values from the attention block. The encoder generates a feature map which is then utilized by the decoder to produce the desired outputs.

#### CNN

As in [32], three convolutional layers and two dense layers make up the convolutional neural network model, with ReLU used as the activation function.

#### LSTM

They are suitable for estimating COVID-19 cases as they have the ability to record time-related dependencies in the data, which is important in time series problems. The LSTM^34-36^ framework was used to forecast COVID-19 transmission due to its ability to handle large amounts of data. The LSTM model used in this study was based on the architecture described in.^32^

#### CNN-LSTM

We blended CNN and LSTM models to forecast COVID-19 cases, like previous research.^32^ The CNN part of the model extracts important information from the data using convolution, generating an embedding. The embedding is then passed to the LSTM, which captures the dependencies over time in the input and makes predictions.

#### Training

The COVID-19 cases forecasting model is trained by using 10 previous daily data points as input. The goal of the algorithm is to forecast the next 4 daily cases based on the input data of 10 previous data points, which serve as the input for the encoder. The decoder will then use this information to generate the next 4 cases. To prevent the model from using future information, a lookahead mask is applied. The transformer model training employs a batch of 64 training examples, a learning rate of 1e-4, and a dropout of 0.05.

## Results and Discussion

Data collected from January 1, 2021, to October 19, 2021, was used to train the DL models. The test data from October 11, 2021, to January 1, 2022, was used to determine the correctness of the models. The performance of the model was evaluated using Root Mean Squared Error (RMSE) and Mean Absolute Error (MAE). As demonstrated in Figure 3, the results show that the Transformer model was able to estimate the daily new cases using the data from the previous day. According to Table 2, the Transformer model proved to be the most effective in forecasting the dissemination of coronavirus cases across different countries, outperforming the CNN and LSTM models. Using a multi-head attention mechanism and positional embeddings, transformers learn by understanding the links between diverse features. The Transformer model’s ability to evaluate just the most relevant features is enhanced through the utilization of probabilistic sparse attention, resulting in more effective training, as seen from the model’s application to different countries. In contrast, LSTMs retain information through a previously computed hidden state due to their sequential processing, while CNNs rely on various kernel sizes to We evaluated the performance of the models by using them to predict COVID-19 cases from January 2nd to 5th 2022. The input for the models was the prediction from the previous day during this forecasting period.

**Table 2:** The comparison of the four COVID-19 forecasting models based on the daily new confirmed cases.

| Country | DL Model | MAE |
| --- | --- | --- |
| India | CNN | 14540 |
|  | LSTM | 28340 |
|  | CNN-LSTM | 20297 |
|  | Informer | 11998 |
| Russia | CNN | 5829 |
|  | LSTM | 2554 |
|  | CNN-LSTM | 3111 |
|  | Informer | 2401 |
| Brazil | CNN | 24659 |
|  | LSTM | 6094 |
|  | CNN-LSTM | 12307 |
|  | Informer | 5368 |

**Figure 3:**
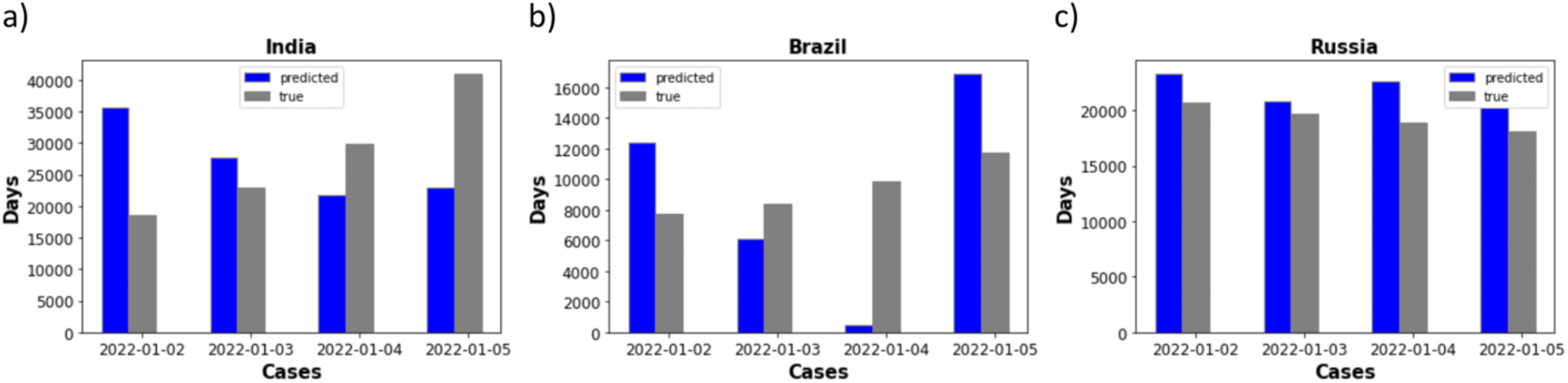
The comparison of the real and predicted daily cases of COVID-19 from January 2nd to 5th, 2022 in India, Brazil, and Russia.

The results of the forecasting of COVID-19 cases from 2nd January 2022 to 5th January 2022 are presented in Figure 4. It is assumed that the regulations in place on the day of forecasting will be the same as the preceding day, which is deemed reasonable. The performance of the Transformer model was found to be the best in all three countries, with the lowest error in India (0.12% with an MAE of 11998 cases), Russia (0.13% with an MAE of 2401 cases), and Brazil (less than 0.1% with an MAE of 5368 cases). Additionally, the Transformer model consistently had a lower RMSE compared to other models, with a difference of 12% over the next-best model (CNN) in India, 4% over the second-best model (LSTM) in Russia, and 4% over the second-best model (LSTM) in Brazil.

**Figure 4:**
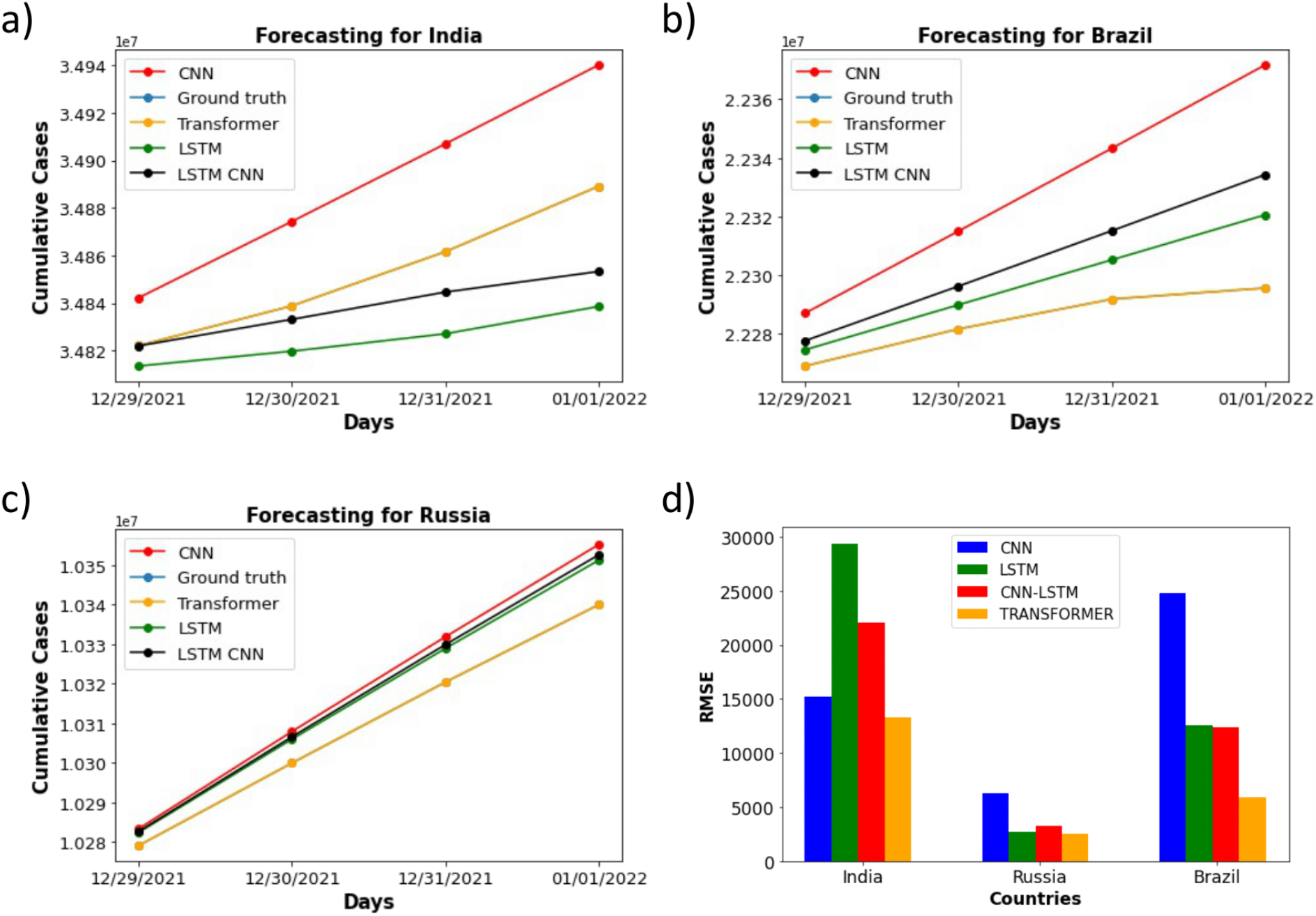
Aggregated COVID-19 cases predicted between 29 December 2021 and 1 January 2022 of (a) India, (b) Brazil, and (c) Russia (d) RMSE results for forecasting models

## Conclusion

To estimate the progression of COVID-19 in three highly impacted nations, a model based on Transformer architecture was developed in this research. This model was compared to three other deep learning models and was found to accurately depict the dissemination of the virus in each nation. The result of using the MAE and RMSE evaluation methods indicates that the Transformer model was the most successful, outperforming CNNs and LSTMs. Our findings suggest that the Transformers can be used to estimate future COVID-19 cases and assist governments in making informed decisions for managing the pandemic. In the future, the model can be further enhanced by incorporating more factors such as variant type, vaccination status, and demographic information. The data and code used can be found at: https://github.com/Saupatil07/Covid-Forecasting-using-deep-learning.

## Data Availability

https://github.com/Saupatil07/Covid-Forecasting-using-deep-learning

## Acknowledgement

This work has been supported by Center for Machine Learning in Health (CMLH) at Carnegie Mellon University and a funding from CMU Mechanical Engineering.

